# Calibration, Uncertainty, and Data Efficiency of Deep Learning Chest Radiograph Classifiers Under Cross-Dataset Distribution Shift: A Multi-Dataset Benchmark

**DOI:** 10.64898/2026.10.14.26365516

**Authors:** Faraz Shamim, Raziya Akhtar Hussain

## Abstract

**Purpose:** To evaluate the external transportability of source-derived probability calibration for chest radiograph deep learning classifiers under distribution shift and determine its dependence on sample size and disease prevalence.

**Materials and Methods:** This retrospective secondary computational analysis of public, de-identified radiographic datasets involved no participant contact or new data collection. Cohort sizes were prespecified by availability without prospective power calculation: 106,686 NIH ChestX-ray14 images (29,323 patients; 1992-2015) for development and internal testing, 20,000 CheXpert images (20,000 patients; October 2002-July 2017), and 14,998 VinBigData images (2018-2020) for external testing. ResNet-18, DenseNet-121, and DeiT-Tiny classifiers for cardiomegaly and pleural effusion were trained across four patient fractions and three seeds (36 fits). Calibrators were fitted exclusively on NIH data. Twelve primary external Brier-score contrasts (temperature-scaled versus raw probabilities) were evaluated using paired cluster-bootstrapping with Holm adjustment.

**Results:** Temperature scaling reduced Brier scores in nine of 12 primary contrasts but worsened pleural effusion calibration on VinBigData across all architectures (Brier increases: ResNet-18, 0.001460 [95% CI: 0.001347, 0.001568]; DenseNet-121, 0.001193 [95% CI: 0.001109, 0.001274]; DeiT-Tiny, 0.000498 [95% CI: 0.000417, 0.000577]; all adjusted *P*=.006). Standardizing class weights to NIH prevalence reversed all CheXpert improvements. Although full versus 10% training improved external AUROC across all comparisons, DeiT-Tiny CheXpert pleural effusion Brier score worsened by 0.007039 despite a 0.032466 AUROC gain. At 80% selective image retention, calibrated ensembles retained only 12.3%-17.2% of VinBigData positive pleural effusion examinations.

**Conclusion:** Source-fitted calibration is not uniformly transportable across health systems, and discrimination gains do not guarantee calibrated probabilities. External evaluation must simultaneously assess probability reliability, prevalence shifts, and retained-case composition.

## 1. Introduction

A chest-radiograph classifier can rank abnormal images above normal images without assigning reliable probabilities. Discrimination describes this ranking, whereas calibration concerns agreement between predicted probabilities and observed event frequencies. Their distinction matters when model outputs are used to prioritize examinations, support decisions, or identify cases requiring review. Calibration is therefore not established by a high area under the receiver-operating-characteristic curve (AUROC) alone [1]. In imbalanced radiographic tasks, apparently favorable summary metrics can also conceal weaknesses affecting the less frequent finding [2].

Post-hoc temperature scaling offers a computationally inexpensive way to adjust probabilities using a held-out labeled cohort [3]. However, its performance after transfer to a different dataset is not guaranteed. Broad uncertainty benchmarks have demonstrated failures of conventional post-hoc calibration under distribution shift [4], and chest-radiograph studies have shown that cross-institutional performance can differ from internal performance [5]. More recently, Roschewitz and colleagues evaluated calibration across natural imaging shifts and found that calibration strategy, pretraining, and the order of calibration and ensembling all affected robustness [6]. Thus, whether source calibration transfers is an empirical question, not a property established once during model development.

Two related issues complicate this evaluation. First, medical labels have a development cost beyond fitting the image classifier: tuning and calibration also require labeled data. A model that performs well with a small training fraction may still depend on a substantial fixed development cohort. Second, probability scores and uncertainty-based selection depend on case mix. A change in the frequency of positive findings can alter aggregate Brier scores, while a low error rate among retained images can reflect preferential retention of negative examinations rather than broad reliability.

We conducted a focused benchmark of two shared chest-radiographic findings across NIH ChestX-ray14, CheXpert, and VinBigData. The primary objective of this benchmark was to quantify the transportability of post-hoc temperature scaling when calibrators are fitted exclusively on source-domain radiographs and evaluated across distinct external health systems. Secondary objectives were to determine how training sample efficiency and calibration cohort size govern probability reliability, and to evaluate how lightweight uncertainty baselines perform under selective prediction. Rather than introducing architectural variations, this study provides a standardized, multi-institutional evaluation of how probability calibration, case-mix variation, and deferred triage interact under distribution shift.

## 2. Materials and Methods

### 2.1 Study Design, Ethics, and Prediction Targets

This retrospective multi-institutional benchmark evaluated image-level classification, probability calibration, and selective triage for cardiomegaly and pleural effusion as independent binary targets. Development and internal testing utilized NIH ChestX-ray14; external testing evaluated CheXpert and VinBigData [7–9].

#### Ethics and Data Governance

This study was a secondary computational analysis of publicly accessible, de-identified radiographic datasets involving no participant contact, recruitment, or new clinical data collection. Ethics approval, consent, and de-identification procedures for the original data collections are described in the respective source publications [7–9].

All model architectures, training fractions, random initialization seeds, calibration routines, decision thresholds, and 12 primary statistical contrasts were locked prior to external evaluation. Model fitting and recalibration were restricted strictly to source-domain data (NIH ChestX-ray14) without access to external test sets. Protocol specifications, split manifests, and reproducible code are available at https://github.com/faraz-shamim/medshift-benchmark.

### 2.2 Cohort Construction, Acquisition Dates, and Reference Standards

Radiographs represented clinical examinations acquired between 1992 and 2020 across three distinct health systems:

1. **NIH ChestX-ray14 (Source Domain; 1992–2015):** Adult radiographs (aged 18–100 years, AP/PA projections) were split 60:10:10:20 into patient-disjoint training, tuning, calibration, and internal-test sets using a fixed seed. Ingestion verified image decodability and intensity variation, collapsing identical intra-patient duplicates to a single instance and purging exact-pixel copies across distinct identifiers.
2. **CheXpert (External Domain 1; October 2002–July 2017):** One frontal radiograph per eligible adult patient (18–100 years) was sampled deterministically independent of findings, capped at 20,000 patients from the public training release [8].
3. **VinBigData / VinDr-CXR (External Domain 2; 2018–2020):** Evaluated all 14,998 publicly distributed PA radiographs from two hospital networks in Vietnam, annotated by multiple independent radiologists [9]. The absence of patient identifiers and demographics in the mirror precluded patient-level clustering or age filtering.

Reference standards followed cohort ascertainment protocols. NIH findings were coded positive if present and negative if absent from text-mined reports. CheXpert explicit positive report mentions were coded positive, explicit negative or absent mentions negative, and uncertain annotations masked per finding. VinBigData findings were coded positive upon consensus agreement by at least two of three independent radiologists (multiple bounding boxes per reader collapsed to one vote). These standards harmonize clinical terminology while capturing differences between institutional NLP extraction and reader consensus.

### 2.3 Image Preprocessing, Model Architectures, and Training Regimens

Radiographs were bilinearly resized to 224 × 224 pixels without cropping, replicated across three channels, and ImageNet-normalized. Augmentation included random rotations (≤ 5°), translations (≤ 2%), and brightness/contrast shifts (≤ 10%). Flips, crops, class rebalancing, label smoothing, MixUp, and CutMix were omitted.

We evaluated three ImageNet-pretrained vision backbones: ResNet-18 (11,177,538 parameters), DenseNet-121 (6,955,906), and DeiT-Tiny (5,524,802) [10–12] (timm identifiers: resnet18.a1_in1k, densenet121.ra_in1k, and deit_tiny_patch16_224.fb_in1k). Each was coupled with a dropout layer (*p* = 0.2) and a two-output linear head.

Models were trained on nested 10%, 25%, 50%, and 100% subsets of source-training patients across three seeds (17, 43, 101; 36 fits), holding tuning and calibration partitions fixed. Training used unweighted binary cross-entropy, AdamW (backbone learning rate 10^−4^, head 10^−3^, weight decay 10^−4^), two warmup epochs, cosine decay, effective batch size 64 (microbatch 32 with gradient accumulation), and 16-bit mixed precision for up to 20 epochs. Checkpoints were selected by tuning-set negative log likelihood (NLL; patience of four epochs after five burn-in epochs; 29.59 cumulative GPU hours).

### 2.4 Calibration Protocols and Uncertainty Quantification

Raw predictions were sigmoid-transformed logits (*z*). Finding-specific temperature scaling estimated scalar *T* > 0 by minimizing NLL on source-calibration images (*p* = *σ*(*z*/*T*)). Positive-slope logistic recalibration estimated *p* = *σ*(*az* + *b*)(*a* > 0) on the same partition. Both preserve ranking. Fits required ≥ 20 positive and negative calibration observations; otherwise, they were marked unestimable. Calibration sample efficiency was tested on nested 10%, 25%, 50%, and 100% calibration-patient subsets using full-training classifiers.

Epistemic uncertainty baselines comprised a 20-sample Monte Carlo dropout pass through the classification head (deterministic backbone) and three-seed probability ensembles [13,14]. Ensemble predictions averaged member probabilities; calibrated ensemble outputs temperature-scaled the logit of pooled mean probabilities. Decision thresholds were calibrated on source-domain predictions to target 90% empirical sensitivity and locked prior to external testing.

### 2.5 Statistical Analysis and Sample Size Justification

#### Sample Size Determination

Cohort sizes were prespecified by repository availability without prospective power calculations: all eligible NIH source examinations (*n* = 106,686), 20,000 CheXpert examinations, and all publicly labeled VinBigData examinations (*n* = 14,998). Non-parametric cluster-bootstrapping quantified precision.

#### Primary Contrast and Hypothesis Testing

The primary estimand was the paired external Brier-score difference between temperature-scaled and raw probabilities at full training data across 12 contrasts (three architectures, two findings, two external domains; negative values favor scaling). Paired differences were averaged across three seeds. Statistical uncertainty was quantified using 2,000 cluster-bootstrap replicates (patient-level for CheXpert and NIH; image-level for VinBigData), generating percentile 95% confidence intervals and two-sided null-centered bootstrap *p*-values. Family-wise error across the 12 primary tests was controlled via Holm-Bonferroni correction (empirical floor: *p* = 1/2,001) [15].

#### Evaluation Metrics

Discrimination was assessed via AUROC and average precision (1,000 cluster-bootstrap replicates of the seed-mean statistic). Probability accuracy was evaluated via Brier score, NLL, 15-bin equal-width/count expected calibration errors (ECE), calibration intercept/slope, reliability diagrams, and a source-prevalence constant baseline. Threshold metrics (sensitivity, specificity, PPV, NPV, F1) were evaluated at source-locked thresholds.

#### Selective Prediction and Sensitivity Analyses

Selective prediction ranked images by predictive entropy, generating error-risk and Brier-risk coverage curves and true-positive retention rates. Sensitivity analyses evaluated CheXpert uncertainty policies (uncertainty-negative, uncertainty-positive, explicit-only), unanimous VinBigData consensus, and demographic/projection subgroups. Prevalence-standardized Brier scores reweighted external class-conditional mean squared errors to NIH calibration prevalence (π): Brier_standardized_ = π × mean[(1 − p)^2^ | Y = 1] + (1 − π) × mean[p^2^ | Y = 0].

This standardization isolates the mathematical impact of target prevalence shift by holding class-conditional error rates constant while removing prevalence-induced metric scaling.

## 3. Results

### 3.1 Cohorts and completed experiments

The final benchmark cohort comprised 141,684 radiographs across three datasets. NIH ChestX-ray14 provided 106,686 images from 29,323 unique patients, partitioned into 64,592 training, 10,538 tuning, 10,227 calibration, and 21,329 internal-test images. External evaluation included 20,000 radiographs from 20,000 CheXpert patients and 14,998 VinBigData radiographs (Table 1).

**Table 1.**
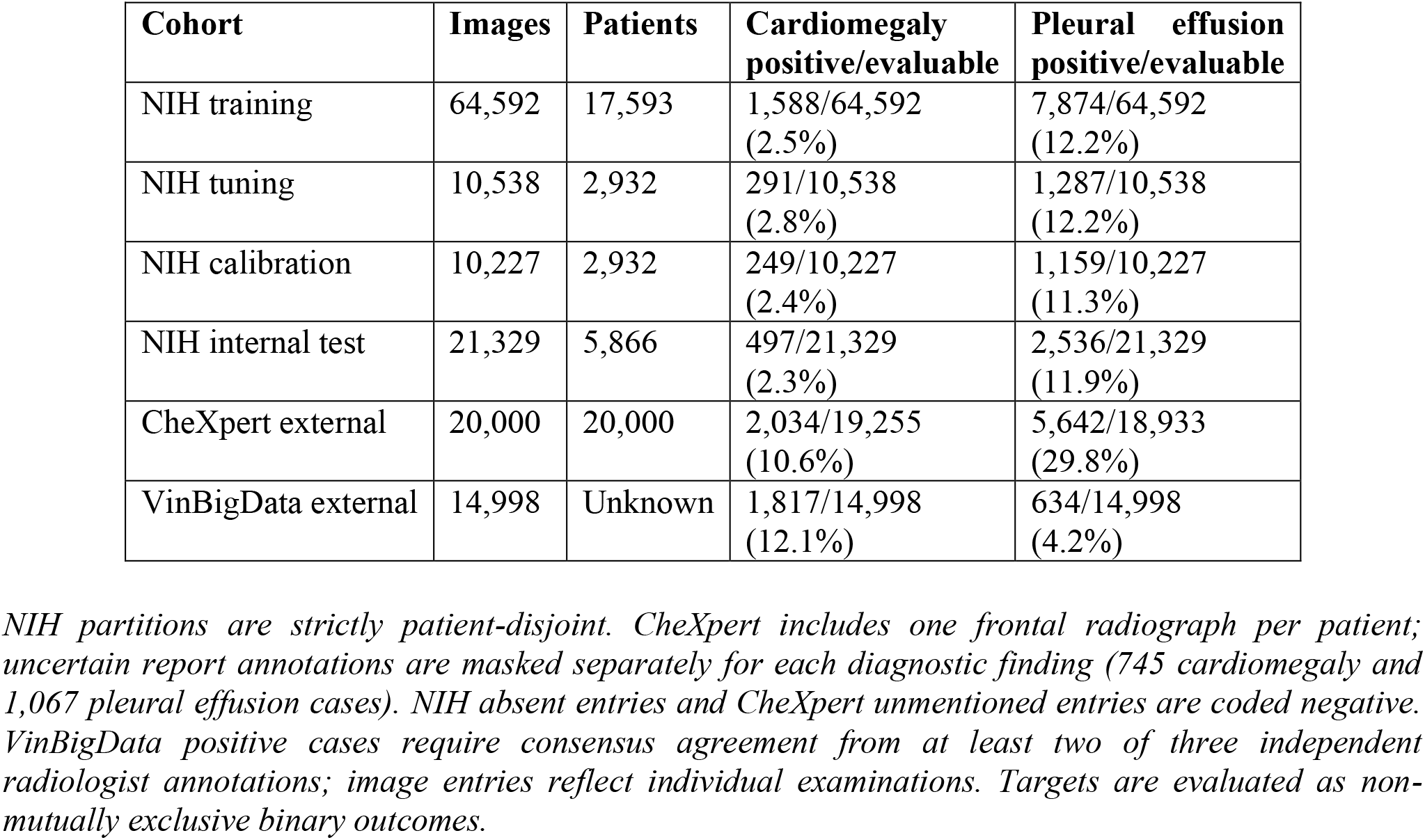
Analyzed cohorts and finding-specific reference labels.

Reference-label prevalence varied substantially across health systems: cardiomegaly prevalence was 2.3% in NIH internal test images, 10.6% in evaluable CheXpert images, and 12.1% in VinBigData; pleural effusion prevalence was 11.9%, 29.8%, and 4.2%, respectively. CheXpert uncertainty masking excluded 745 cardiomegaly and 1,067 pleural effusion labels, leaving 19,255 and 18,933 evaluable examinations. Median patient age was 49 years in NIH and 62 years in CheXpert; demographic metadata were unavailable in VinBigData.

All 36 planned training configurations completed successfully. The 10% patient subsets contained 1,760 patients (6,264–6,852 images), whereas full training encompassed 17,593 patients (64,592 images). Including fixed tuning and calibration partitions (20,765 images), the smallest training budget utilized 27,029–27,617 total labeled images (approximately 32% of the 85,357 images at full development). Individual runs converged within 6–14 epochs (319 epochs cumulatively; 29.59 total GPU hours across parallel nodes).

### 3.2 Internal and external performance

At full training, mean raw cardiomegaly AUROC ranged from 0.884 to 0.892 internally, compared with 0.773–0.800 on CheXpert and 0.852–0.872 on VinBigData. Pleural-effusion AUROC was 0.871–0.883 internally, 0.847–0.862 on CheXpert, and 0.898–0.916 on VinBigData. External discrimination did not uniformly decline: pleural-effusion ranking on VinBigData exceeded internal performance across all architectures (Table 2).

**Table 2.** Full-training discrimination and probability evaluation.

| **Cohort / finding** | **Model** | **Raw AUROC**<br>**(95% CI)** | **Brier**<br>**raw** | **Brier**<br>**TS** | **ECE**<br>**raw → TS** |
| --- | --- | --- | --- | --- | --- |
| NIH<br>Cardiomegaly | ResNet-18 | 0.884 (0.868 to 0.901) | 0.02012 | 0.02015 | 0.004 → 0.005 |
| NIH<br>Cardiomegaly | DenseNet-121 | 0.892 (0.875 to 0.908) | 0.02036 | 0.02033 | 0.006 → 0.005 |
| NIH<br>Cardiomegaly | DeiT-Tiny | 0.891 (0.874 to 0.907) | 0.02068 | 0.02065 | 0.008 → 0.007 |
| CheXpert<br>Cardiomegaly | ResNet-18 | 0.773 (0.764 to 0.783) | 0.09326 | 0.09263 | 0.081 → 0.079 |
| CheXpert<br>Cardiomegaly | DenseNet-121 | 0.800 (0.790 to 0.809) | 0.08723 | 0.08686 | 0.071 → 0.069 |
| CheXpert<br>Cardiomegaly | DeiT-Tiny | 0.800 (0.791 to 0.809) | 0.08639 | 0.08573 | 0.066 → 0.064 |
| VinBigData<br>Cardiomegaly | ResNet-18 | 0.863 (0.856 to 0.870) | 0.09991 | 0.09903 | 0.092 → 0.090 |
| VinBigData<br>Cardiomegaly | DenseNet-121 | 0.852 (0.845 to 0.859) | 0.10190 | 0.10145 | 0.093 → 0.092 |
| VinBigData<br>Cardiomegaly | DeiT-Tiny | 0.872 (0.865 to 0.879) | 0.09635 | 0.09542 | 0.087 → 0.085 |
| NIH<br>Pleural effusion | ResNet-18 | 0.871 (0.861 to 0.881) | 0.07747 | 0.07750 | 0.011 → 0.009 |
| NIH<br>Pleural effusion | DenseNet-121 | 0.883 (0.874 to 0.893) | 0.07516 | 0.07530 | 0.018 → 0.017 |
| NIH<br>Pleural effusion | DeiT-Tiny | 0.878 (0.868 to 0.888) | 0.07649 | 0.07626 | 0.014 → 0.010 |
| CheXpert<br>Pleural effusion | ResNet-18 | 0.847 (0.841 to 0.852) | 0.18248 | 0.17636 | 0.163 → 0.145 |
| CheXpert<br>Pleural effusion | DenseNet-121 | 0.862 (0.857 to 0.867) | 0.16274 | 0.15890 | 0.137 → 0.124 |
| CheXpert<br>Pleural effusion | DeiT-Tiny | 0.854 (0.849 to 0.859) | 0.16702 | 0.16302 | 0.134 → 0.121 |
| VinBigData<br>Pleural effusion | ResNet-18 | 0.898 (0.884 to 0.910) | 0.02819 | 0.02965 | 0.031 → 0.045 |
| VinBigData<br>Pleural effusion | DenseNet-121 | 0.916 (0.903 to 0.928) | 0.02614 | 0.02733 | 0.036 → 0.046 |
| VinBigData<br>Pleural effusion | DeiT-Tiny | 0.915 (0.903 to 0.928) | 0.02496 | 0.02546 | 0.024 → 0.033 |

*TS = source-fitted temperature scaling; ECE = 15-bin equal-width expected calibration error. Metrics are means across three fixed trained seeds, not metrics of averaged probabilities. AUROC confidence intervals use 1,000 cluster-bootstrap replicates and condition on the fitted models. TS preserves AUROC. Brier and ECE are unitless and lower is better; Brier is not a pure calibration measure. Native-prevalence scores from different cohorts should not be compared without their reference-positive fractions. Seed SDs, additional metrics, and method-specific intervals are available in supplementary data.*

Probability evaluation demonstrated marked transfer degradation. Raw cardiomegaly ECE was 0.004– 0.008 internally, 0.066–0.081 on CheXpert, and 0.087–0.093 on VinBigData. Raw CheXpert pleural effusion Brier scores were 0.162742–0.182478 versus 0.024963–0.028188 on VinBigData, reflecting differing label prevalences. Reliability diagrams confirmed persistent mismatch after source calibration, including risk underprediction across intermediate CheXpert pleural effusion probabilities (Figure 2).

### 3.4 Primary temperature-scaling comparisons

Under native target prevalences, temperature scaling improved seed-mean Brier scores in nine of 12 primary contrasts. For cardiomegaly, absolute reductions were modest (0.000369–0.000668 on CheXpert; 0.000447–0.000930 on VinBigData). Pleural effusion improvements on CheXpert were larger: 0.006120 for ResNet-18, 0.003841 for DenseNet-121, and 0.004010 for DeiT-Tiny (Table 3; Figure 1).

**Table 3.** Primary external Brier-score contrasts at full training.

| **Cohort / finding** | **Model** | **TS – raw** | **95% CI** | **Holm p** |
| --- | --- | --- | --- | --- |
| CheXpert<br>Cardiomegaly | ResNet-18 | -0.000638 | -0.000681 to -0.000599 | 0.006 |
| CheXpert<br>Cardiomegaly | DenseNet-121 | -0.000369 | -0.000397 to -0.000342 | 0.006 |
| CheXpert<br>Cardiomegaly | DeiT-Tiny | -0.000668 | -0.000713 to -0.000626 | 0.006 |
| VinBigData<br>Cardiomegaly | ResNet-18 | -0.000878 | -0.000932 to -0.000827 | 0.006 |
| VinBigData<br>Cardiomegaly | DenseNet-121 | -0.000447 | -0.000479 to -0.000418 | 0.006 |
| VinBigData<br>Cardiomegaly | DeiT-Tiny | -0.000930 | -0.000983 to -0.000877 | 0.006 |
| CheXpert<br>Pleural effusion | ResNet-18 | -0.006120 | -0.006398 to -0.005850 | 0.006 |
| CheXpert<br>Pleural effusion | DenseNet-121 | -0.003841 | -0.004039 to -0.003643 | 0.006 |
| CheXpert<br>Pleural effusion | DeiT-Tiny | -0.004010 | -0.004217 to -0.003817 | 0.006 |
| VinBigData<br>Pleural effusion | ResNet-18 | +0.001460 | +0.001347 to +0.001568 | 0.006 |
| VinBigData<br>Pleural effusion | DenseNet-121 | +0.001193 | +0.001109 to +0.001274 | 0.006 |
| VinBigData<br>Pleural effusion | DeiT-Tiny | +0.000498 | +0.000417 to +0.000577 | 0.006 |

*Negative values favor TS. Paired per-image losses were averaged across three fixed seeds before 2,000 cluster-bootstrap replicates (patients for CheXpert; images for VinBigData). Intervals are pointwise and conditional on fitted models and calibrators. All unadjusted approximate bootstrap p values reached the finite-resolution floor of 1/2,001; Holm adjustment gives $12/2,001 = 0.005997$ , displayed as 0.006. These equal p values do not imply equal effects or simultaneous confidence intervals.*

**Figure 1.**
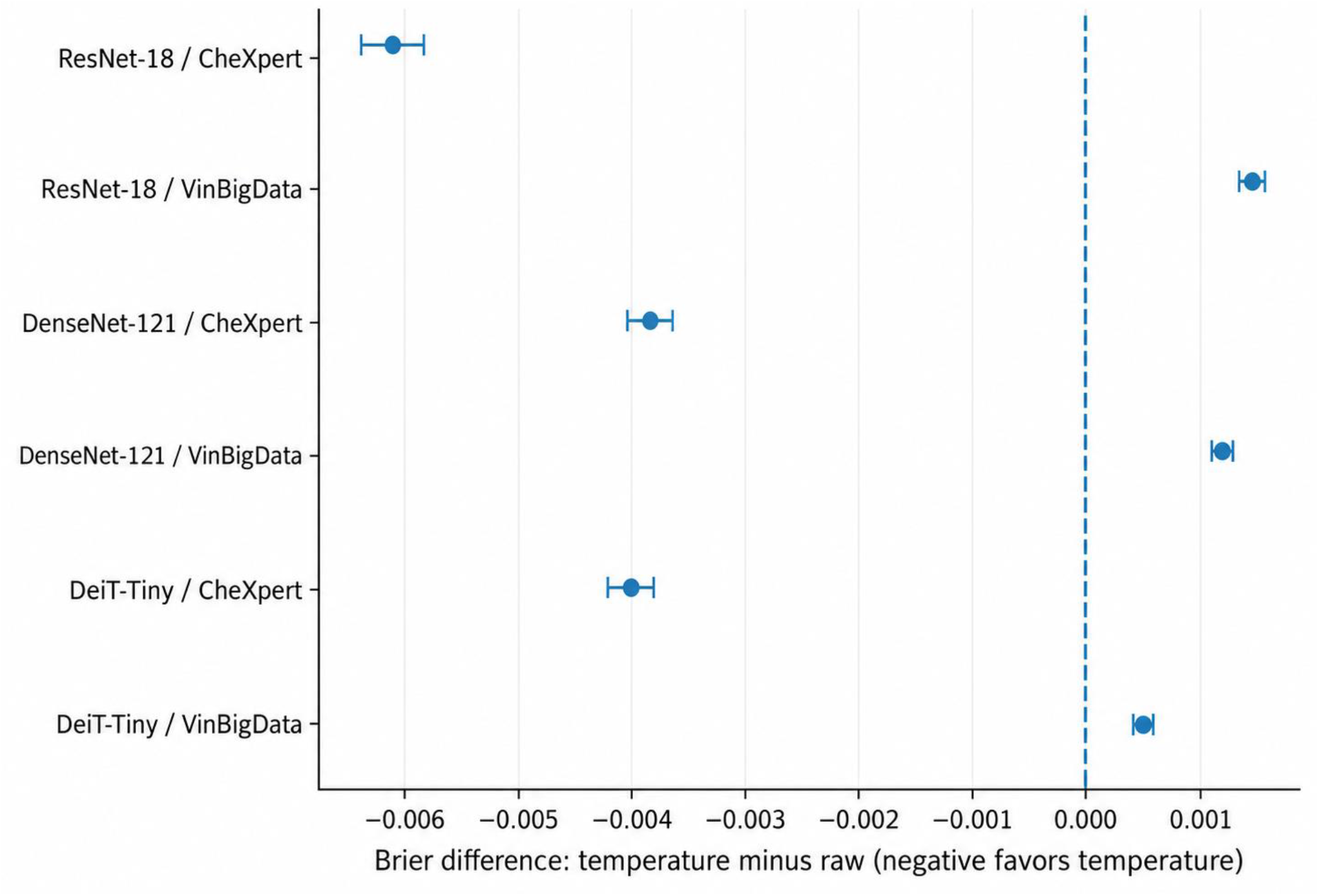
Primary paired Brier-score contrasts for pleural effusion following source-domain temperature scaling. External Brier-score differences (Brier_TS_ - Brier_raw_) across ResNet-18, DenseNet-121, and DeiT-Tiny architectures trained at full sample size. Negative values indicate improved probability accuracy under temperature scaling. Source calibration consistently improves native-prevalence probability accuracy on CheXpert while degrading accuracy on VinBigData across all three architectures. Points indicate seed-mean estimates; error bars denote pointwise 95% confidence intervals derived from 2,000 paired cluster-bootstrap replicates conditional on fitted model checkpoints. Numerical values and cardiomegaly comparisons are detailed in Table 3; prevalence-standardized comparisons are presented in Table 4.

**Figure 2.**
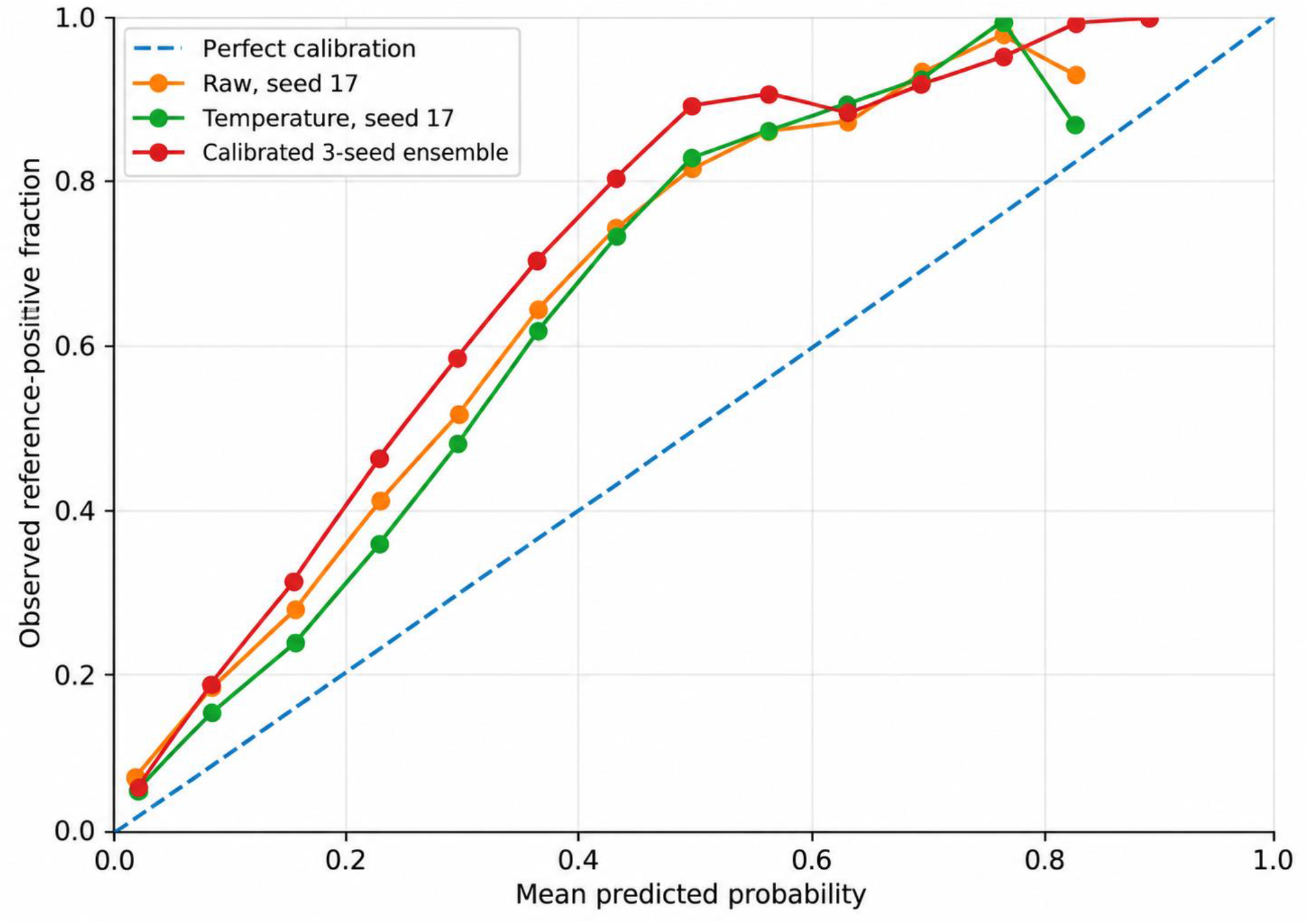
External reliability diagrams for pleural effusion prediction on CheXpert. Observed reference-positive event frequency plotted against mean predicted probability across 15 equal-width bins for DenseNet-121 models trained at full sample size. The diagonal dashed line denotes perfect calibration. Curves illustrate uncalibrated predictions (raw, seed 17), source temperature-scaled predictions (seed 17), and the pooled three-seed probability ensemble temperature-scaled after pooling. Source-fitted calibrators systematically underestimate risk across the intermediate probability range (0.2– 0.6) under target distribution shift.

Conversely, temperature scaling systematically degraded VinBigData pleural effusion calibration across all architectures. Brier scores increased from 0.028188 to 0.029648 for ResNet-18, from 0.026135 to 0.027328 for DenseNet-121, and from 0.024963 to 0.025461 for DeiT-Tiny. Paired differences and 95% confidence intervals were +0.001460 (0.001347–0.001568), +0.001193 (0.001109–0.001274), and +0.000498 (0.000417–0.000577), respectively (all Holm-adjusted *P* = .006). VinBigData pleural effusion NLL and ECE similarly degraded, while AUROC was preserved.

**Table 4.**
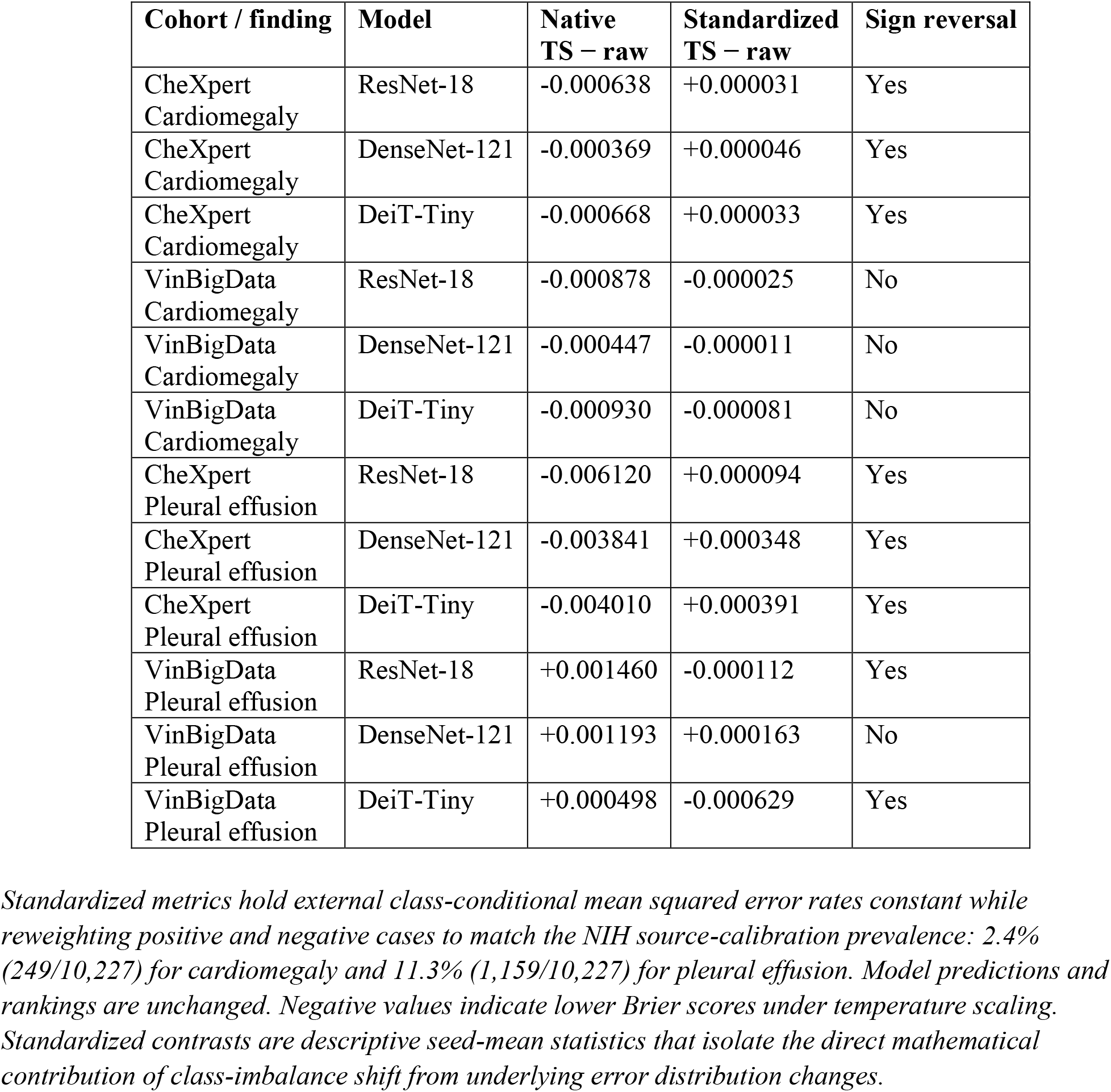
Sensitivity of the Brier contrast to prevalence standardization.

Individual fits showed seed sensitivity for cardiomegaly (two DenseNet-121 seeds and one DeiT-Tiny seed showed Brier degradations), whereas pleural effusion fits uniformly improved on CheXpert and degraded on VinBigData.

### 3.4 Prevalence and label-policy sensitivity

Standardizing external class-conditional error distributions to source NIH calibration prevalence substantially altered primary outcomes (Table 4). When class weights were equalized to source prevalence, all six CheXpert Brier improvements reversed into slight deteriorations (+0.000031 to +0.000391). For VinBigData pleural effusion, standardization reversed ResNet-18 and DeiT-Tiny degradations into net improvements (−0.000112 and −0.000629), while DenseNet-121 remained marginally worse (+0.000163). VinBigData cardiomegaly improvements remained negative but attenuated.

These standardized contrasts demonstrate that shifts in positive-to-negative outcome ratios strongly govern aggregate probability scores independent of underlying feature representation. Native-prevalence temperature scaling patterns persisted under alternative label policies. Restricting CheXpert evaluation to explicit binary mentions reduced sample sizes to 3,371 cardiomegaly and 9,850 pleural effusion cases (positive fractions: 60.3% and 57.3%). Unanimous three-reader consensus on VinBigData retained 13,995 cardiomegaly and 14,405 pleural effusion cases (Supplementary Table S7).

### 3.5 Training-data and calibration-data efficiency

Scaling training data from 10% to 100% improved mean temperature-scaled AUROC across all 12 configurations (gains of 0.032–0.158) and improved Brier scores in 11 configurations. However, DeiT-Tiny pleural effusion on CheXpert revealed discordance between ranking and calibration: expanding training volume increased AUROC from 0.821738 to 0.854204 (Δ = +0.032466; 95% CI, 0.028858–0.035952), yet Brier score degraded from 0.155976 to 0.163015 (Δ = +0.007039; 95% CI, 0.005441– 0.008647). Learning trajectories showed non-monotonic behavior across intermediate subsets, confirming that discrimination gains do not consistently track probability accuracy (Figure 3).

**Figure 3.**
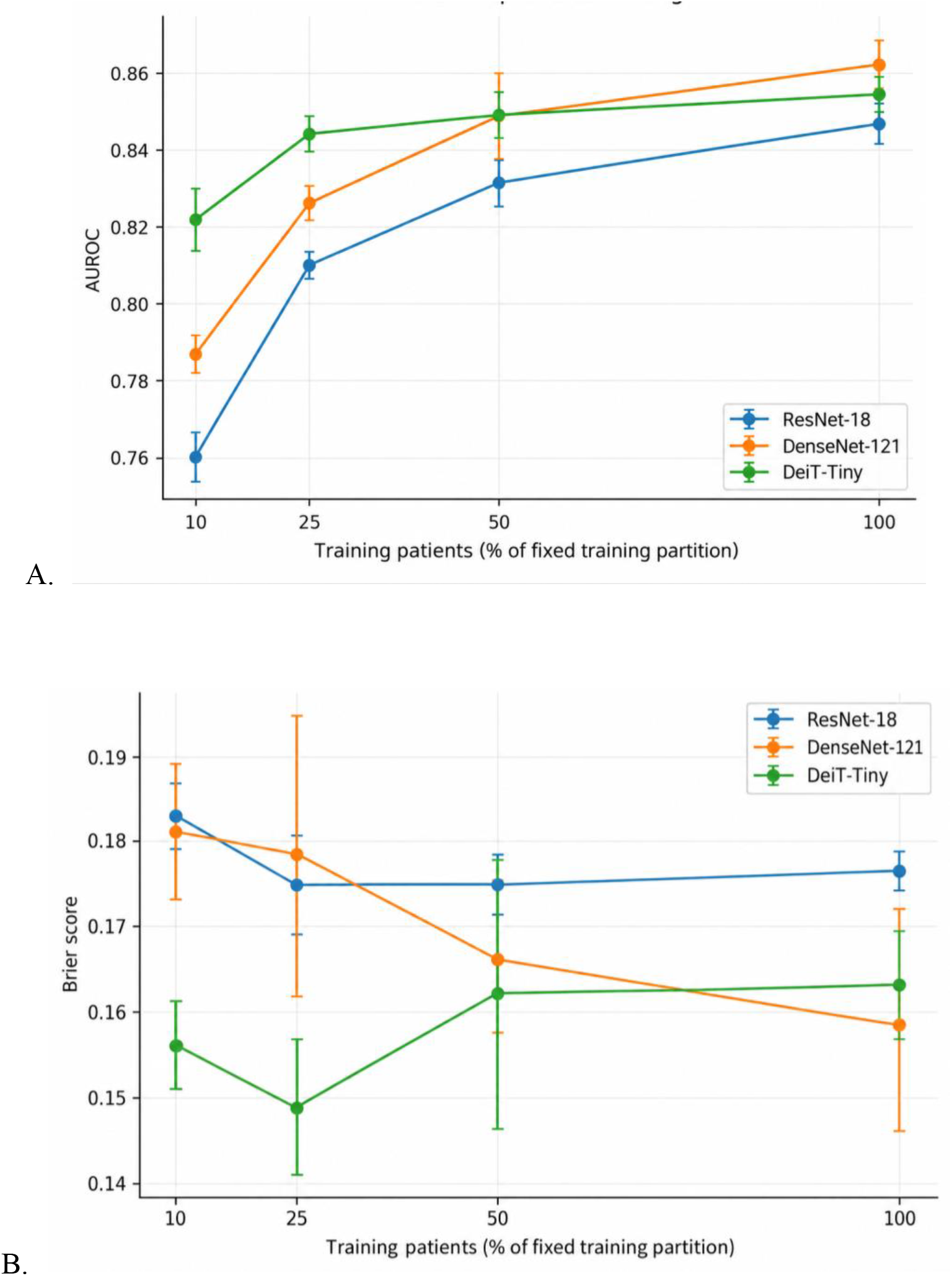
Divergence between discrimination and probability accuracy across training-data budgets. Learning trajectories for pleural effusion prediction on CheXpert following source temperature scaling across 10%, 25%, 50%, and 100% patient training fractions. **(A)** Area under the ROC curve (AUROC) and **(B)** Brier score. Points and error bars denote means and ±1 standard deviation across three random initialization seeds. Source tuning and calibration partitions were held constant across all training fractions. For DeiT-Tiny pleural effusion on CheXpert, AUROC improved monotonically with increasing training data, whereas calibrated Brier score followed a non-monotonic trajectory and was ultimately worse at 100% training than at 10%.

In the calibration-budget analysis, seed 43 at 10% calibration (294 patients, 955 images) lacked the required 20 cardiomegaly events, leaving six fits unestimable across architectures. The remaining seeds met the threshold, allowing two-run averages for 10% cardiomegaly. All eligible calibration routines converged normally, highlighting that fixed patient fractions do not guarantee sufficient event counts for low-prevalence findings.

### 3.6 Uncertainty baselines and retained-case composition

Uncalibrated three-seed probability ensembles yielded lower external Brier scores and higher AUROC values than individual constituent models across all 12 comparisons (e.g., VinBigData pleural effusion AUROCs: 0.907 for ResNet-18, 0.925 for DenseNet-121, and 0.927 for DeiT-Tiny). While Brier reductions are mathematically expected when averaging squared probability errors, post-hoc calibration of pooled ensemble logits produced divergent results across domains. Head-only Monte Carlo dropout yielded negligible adjustments (∣ ΔBrier ∣< 0.00024).

Entropy-based selective prediction revealed pronounced asymmetry in retained-case composition under distribution shift. For VinBigData pleural effusion, selecting the 80% most confident examinations with calibrated ensembles retained only 17.2% of true-positive cases for ResNet-18, 12.9% for DenseNet-121, and 12.3% for DeiT-Tiny (Figure 4). While error rates within the retained cohort were markedly reduced, this was achieved by deferring over 80% of abnormal cases rather than maintaining uniform confidence across classes.

**Figure 4.**
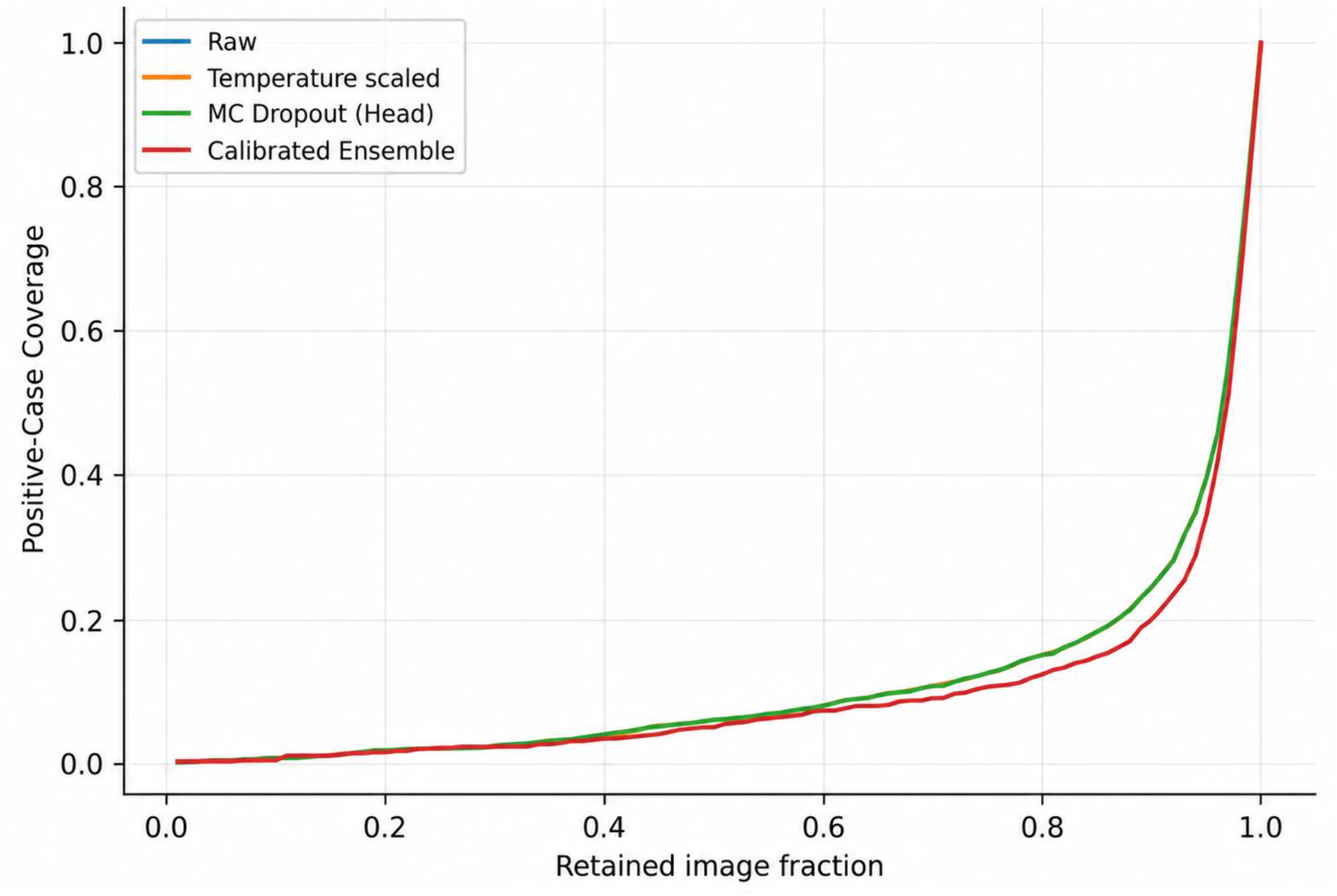
True-positive retention during entropy-based selective evaluation for DeiT-Tiny pleural-effusion detection on VinBigData. Positive-case coverage, defined as the proportion of all reference-positive examinations retained, plotted as a function of total retained image fraction under entropy-based uncertainty ranking with source-locked operating thresholds. At 80% overall image retention, the calibrated three-seed probability ensemble retains only 12.3% of true-positive examinations, directing 87.7% of abnormal cases to the uncertainty pool. Curves compare uncalibrated, temperature-scaled, Monte Carlo dropout, and calibrated ensemble inference.

## 4. Discussion

This benchmark demonstrates that source-fitted calibration does not uniformly transfer across clinical datasets. While temperature scaling improved mean Brier scores in nine of 12 native-prevalence comparisons, it systematically degraded pleural effusion calibration on VinBigData across all architectures without altering AUROC. Crucially, prevalence standardization reversed several aggregate outcomes. The principal finding is therefore not simply that calibration deteriorated in one cohort, but that its apparent transportability varied with reference-label prevalence, target case mix, and error weighting in addition to model discrimination.

These findings corroborate prior machine learning benchmarks showing that probability calibration degrades independently of discrimination under dataset shift [4,6], extending the class-imbalance insights of Mosquera et al. [2] to multi-institutional radiography. By evaluating calibration alongside classifier-training and calibration-label budgets across three distinct backbones within a unified protocol, our results indicate that calibration transportability appears to vary with target-domain prevalence, task-specific probability distributions, and dataset shift, rather than being determined by neural architecture family alone.

Prevalence differences explain the divergent Brier score behaviors. NIH calibration pleural effusion prevalence was 11.3%, compared with 29.8% in CheXpert and 4.2% in VinBigData. A post-hoc recalibration mapping can reduce squared errors among positive cases while increasing them among negatives; aggregate Brier score changes reflect their relative weighting. While prevalence standardization cannot isolate biological case mix from differences in institutional acquisition, projection, or annotation, it demonstrates how class proportions drive summary reliability metrics. Reporting native and standardized scores together clarifies performance across differing target populations.

Training volume and probability accuracy exhibited discordant trajectories. For DeiT-Tiny pleural effusion on CheXpert, expanding source-training data increased AUROC while progressively deteriorating post-hoc calibrated Brier scores. Evaluating data efficiency through discrimination alone can therefore conceal severe probability degradation. Comprehensive assessments of data efficiency must specify the target utility metric and account for the fixed annotation costs of tuning and calibration cohorts alongside base training volumes.

The calibration-budget experiment offers a complementary practical observation: fixed patient fractions do not guarantee sufficient positive event counts for rare findings. At 10% calibration, cardiomegaly calibrators were estimable for two seeds but failed in the third due to insufficient positive events. This underscores the necessity of reporting absolute event frequencies and optimization failures rather than assuming that uniform patient sampling provides adequate calibration data across disease targets.

The uncertainty baselines demonstrated practical trade-offs. Probability ensembling consistently reduced Brier scores through variance reduction under squared loss, whereas head-only Monte Carlo dropout produced minimal adjustments [13,14]. Importantly, strictly positive temperature scaling preserves logit ordering in binary classification, leaving entropy-based risk–coverage curves identical up to numerical precision. Unchanged selective-prediction curves following temperature scaling thus reflect monotonic rank preservation rather than invariant or improved uncertainty representations.

The selective prediction analysis exposes a critical operational blind spot. Under distribution shift, models achieved low retained error rates by deferring over 80% of abnormal examinations to the uncertainty pool. A system that retains predominantly negative examinations functions as a rule-out screening tool rather than a comprehensive classifier, returning subtle pathology to the radiologist. Benchmarks evaluating uncertainty-based triage must report positive-case retention rates alongside overall coverage to clarify real-world clinical utility.

Several methodological boundaries define the analytical scope of this benchmark. First, evaluation was restricted to two thoracic findings across three public radiography cohorts; calibration transportability and selective-deferral dynamics may differ for subtle focal lesions, multi-label classifications, alternative imaging modalities, or vision-language foundation models. Second, reference standards relied on distinct ascertainment mechanisms—automated natural-language-processed hospital reports (NIH, CheXpert) versus localized multi-radiologist consensus votes (VinBigData)—which remain inherently coupled with institutional imaging shifts. Third, radiographs were uniformly downsampled to 224 × 224 pixels without preserving native anatomical aspect ratios to evaluate standard vision backbones, which may attenuate fine radiographic features. Finally, the absence of verified patient identifiers and demographic metadata in the distributed VinBigData mirror precluded cross-institutional patient-overlap audits and necessitated image-level rather than patient-level cluster-bootstrapping for that cohort.

Analytical precision, experimental design, and computational reproducibility also contextualize these findings. Bootstrap confidence intervals quantify test-cohort sampling variability conditional on fixed model checkpoints and calibrators, rather than uncertainty from development-population resampling. While three random seeds captured initialization variance—demonstrating seed sensitivity for certain cardiomegaly calibration effects—broader sweeps would be needed to characterize complete retraining distributions. Models were trained under a unified optimization recipe rather than architecture-specific exhaustive hyperparameter tuning to maintain standardized comparative transfer conditions. Subgroup analyses were similarly constrained by incomplete demographic descriptors in external cohorts, and GPU execution across non-deterministic CUDA operations introduces subtle numerical variance. In accordance with CLAIM 2024 guidelines [16], complete software pipelines, execution manifests, and random seeds are provided to ensure transparent provenance and methodological reproducibility, though prospective clinical trials remain necessary to evaluate real-world workflow integration and downstream patient outcomes.

## 5. Conclusions

In this multi-dataset chest radiography benchmark, source-domain temperature scaling exhibited dataset-dependent transportability, improving probability accuracy on CheXpert while systematically degrading calibration on VinBigData for pleural effusion. These outcomes were highly sensitive to target-domain case mix: standardizing evaluation to source prevalence reversed calibration improvements on CheXpert and mitigated degradations on VinBigData. Furthermore, training-data volume produced divergent trajectories between discrimination and calibration, and uncertainty-based deferral achieved low error rates primarily by excluding positive pathology. Robust external evaluation of medical image classifiers requires simultaneous reporting of discrimination, calibrated probability metrics, reference-label prevalences, calibration-event frequencies, and positive-case retention profiles.

## Data Availability

The complete benchmarking protocol, patient split manifests, model training scripts, and calibration evaluation code are publicly available on GitHub at:

https://github.com/faraz-shamim/medshift-benchmark

## Data and Code Availability

All raw image datasets are accessible from their respective hosting providers (NIH ChestX-ray14 via the NIH Clinical Center, CheXpert v1.0 via Stanford AIMI, and VinDr-CXR via PhysioNet). The complete benchmarking protocol, patient split manifests, model training scripts, and calibration evaluation code are publicly available on GitHub at: https://github.com/faraz-shamim/medshift-benchmark

